# Pixaire1: Evaluation of automated chronic wound surface measurement systems

**DOI:** 10.64898/2026.03.30.26344793

**Authors:** Guillaume Maxant, Carine Mori, Thibault Maxant, Anne-Claire Bertaux

## Abstract

**Purpose:** To evaluate two smartphone-based methods for measuring the surface area of chronic wounds using : Woundtrack (semi-automated measurement: WT) and Woundsize (automated measurement: WS), and comparing them with the reference technique: digitized planimetry (PL).

**Population and methods:** Pixaire 1 is an open-label, single-center study involving 42 patients, from May to June 2023.

Wound surfaces were measured using the three methods by two independent experts. We realized a four steps statistical analysis: multivariate analysis of variance; correlation between the two experts (*precision*); agreement between the two evaluated methods and the reference (*accuracy*); analysis of non-conformities (differences of more than 20% in absolute values compared with the PL measurement) in a subset of wound less than 8 cm*≤*^2^.

**Results:** Of the 42 patients, 6 were excluded from the statistical analysis (multiplanar wound: 4; difficult edge delineation: 2).

We found no difference in multivariate analysis

We showed excellent agreement (ICC *>* 0, 9) of repeated measures (precision) for all three protocols. We also demonstrated excellent agreement (ICC *>* 0, 9) between WT and WS measurements versus PL (accuracy). However, accuracy and precision were better for WT than for WS.

Analysis of non-conformities in small areas wounds showed no difference in variance and distribution between WT and PL, and showed a significant difference between WS and PL.

**Conclusion:** Woundtrack is close to Digitized Planimetry, in terms of precision (reproductibility of the measure) and accuracy (correlation of measures with digitized planimetry). Despite the existence of non-conformities in small wounds, WT does not significantly differ of PL in this subset. WT should be considered as an effective method to measure the area of the wound, similar to PL, with a real benefit in implementation in current care setting (easy to realize, less time consuming).

Woundsize showed less consistent results, despite a reliability and an accuracy that remains good. Its integration in a ’Algorithm: propose then Clinician: correct and validate’ procedure seems most efficient way to implement such methods.

## Introduction

### Epidemiology of the chronic wound

Chronic wounds, defined as any loss of skin substance that has not healed after 4 to 6 weeks, are dominated by three etiologies: diabetic foot ulcers (DFU), varicose ulcers (VU) and pressure-associated ulcers (PAU). In any case, healing is a process that involves several mechanisms which may occur simultaneously on the same wound. The clinical evolution of the wound is therefore described in four distinct phases: *detersion*, in which inflammatory processes elimintae the non-viable tissues; *granulation* or proliferative phase, in which fibroblasts proliferation fills the defect and creates a wound bed ; *epidermization*, which involves the coverage of the wound surface by the epidermis, from the edges of the wound; and *remodeling*.

This process can be prolonged or even stopped by several factors, essentially infection, continued pressure, arterial or venous insufficiency, that led to chronicisation.

It has a major and growing impact on the healthcare system, in terms of prevalence and cost. Zhang ([27]) estimates the prevalence of DFU at 6.3% of the diabetic patients. VLU represents a cumulative lifetime risk ranging from 1 to 1.8% [21] affecting 1% of the adults population and 3.6% of the population older than 65 years. Incidence of hospital-acquired PU may be up to 8.4% [17]. On the financial side, weekly cost of VLU ranges froms 214 to 294 dollars in Australia [1].

### Quantifying the chronic wound kinetics

Measuring the surface area of a chronic wound is a fundamental aspect of its follow-up, which is unfortunately rarely used in clinical practice regarding to the time and/or specific devices required to realize the measurement. It provides a quantified assessment of the wound’s evolution enabling a precise adaptation of the care.

Progression kinetics at the time of initial management are therefore a key factor in the prognosis of chronic wounds, as emphasized by Cardinal [4]. In particular, they enable a distinction to be made between refractory wounds, where progress is blocked by local mechanisms (biofilm for example) and would require more invasive management. The use of wound surface measurement methodes is therefore essential to optimize the wound management. Ideally, this method should :

- Reliable (Accuracy: correlation of the measurement with the reference measurement),
- Reproducible over time and between operators (Precision: correlation of measurements taken by different operators) and
- Easy to access, with the use of a user-friendly interface.

### Methods available

The methods available are classified into three categories by Wu [26]. Full contact methods require the placement of elements on the wound (tracing paper or transparency, local trimming elements). Partial contact techniques enable measurement by placing a reference marker in contact with the wound. Non-contact methods enable 3D reconstruction of the wound and therefore measurement without the need for reference markers.

Contact methods consists in manual planimetry and digitized planimetry ([15], [3]), and Visitrak ([22], [12], [9]),

The partial contact methods, implying marker placement are essentially represented by:

- ImitoMeasure [2], [7],
- Silhouette [20], [12],
- Tissue Analytics [8],
- NDKare [14],
- VeVMD and
- Non-dedicated solutions: ImageJ, AutoCAD.

Non-contact methods implies specific capture devices or methods:

- C4W (3D structure sensor) [5],
- WoundVue (Infrared Projector and dual infrared camera),
- 3DWamCamera (projector and 3 camera) [25], [12] and
- Photogrammetry: LifeViz3D, and associated methods (Neural Radiance Field: NeRF, 3D Gaussian Splatting),

We consider digitized planimetry (acetate tracing + measurement using ImageJ software) as the reference method. Lagan [15] and Bilgin [3] showed that acetate tracing was superior to the other conventional methods. Jeffcoates [11] has demonstrated the reproductibility of digitized planimetry, which involves measuring the traced surface using ImageJ.

### WoundTrack and Woundsize

Pixacare solution offers a tool for classification (photo library) and remote monitoring of chronic wounds, . It is based on an interface that can be used directly on mobile devices (smartphones, tablets). Within this framework, it has developed a tool for measuring the wound surface by partial contact, based on two methods which differ according to the process employed for segmenting the wound.

Woundtrack measures the surface area after the operator has delimited the wound edges on the phone’s touch screen.

Woundsize incorporates a deep learning algorithm, trained to automatically delimit the edges of the wound, from which the surface area is measured. Here’s the English translation:

The Pixacare solution offers a classification tool (photo library) and remote monitoring for chronic wounds as we previously shown in [18]. It is based on acquiring photographs and clinical data (called *sequences*) from a smartphone, and organizing them in a patient file stored in a secure cloud. The acquisition is designed to be entirely managed on the smartphone, so that it can be performed in just a few minutes at the patient’s bedside, whether at home, during consultation, or during hospitalization. This method allows access to monitoring data for all caregivers to whom the patient has provided the access key, via a QR code.

Above this photo library, a measurement overlay for surface area and granulation tissue proportion is applied. In this context, placing a normalization sticker (for distance and colorimetry) in the wound plane is necessary. Wound analysis is then accessible, after manual outlining by the clinician (Wountrack 1 algorithm) or automated outlining using a computer vision algorithm (Woudsize). For more detail, a video describing the acquisition and manual outlining is provided as supplementary material, as well as a download link for the application.

This clinical trial (PIXAIRE-1) evaluates the performance of these two algorithms, with a view to clinical validation. Digitized planimetry will be used as the reference method to assess the measurement accuracy. Independent measurements by two operators will be used to assess the accuracy of the methods.

## Population and Methods

### Pixaire 1

Pixaire 1 is a single-center, open-label, cross sectional study. It consisted in the evaluation of three different algorithms, Woundtrack (wound measurement after segmentation of the wound by the care practitioner), Woundsize (wound measurement after automatized segmentation). A third studied algorithm, Woundtype (characterization of the tissues in the wound) will not be exploited in this article.

These two algorithms were compared to several methods of measurement of the size of the chronic wound, such as expert’s eye evaluation, ruler measurement, digitized planimetry.

The study was conducted in the Vascular Surgery Department of Haguenau’s hospital (France), from May to June 2023.

The patients treated in the department, presenting with a wound requiring directed healing of any kind, between 4 and 150 cm^2^ were considered to the inclusion. Exclusion criteria consisted in multiple wounds; wounds that couldn’t be taken in a single photo; presence of elements (tatoo, birthmark) that would make the patient identifiable. Patients was informed of the study and had to consent to take part of it.

This study was the first formalized evaluation of those algorithms in a real-life setting. The power of the test was assessed on the basis of data from non-formal pre-clinical studies carried out previously. With a bilateral approach, an alpha risk of 5%, a performance (concordance = code 1) estimated at 80% can be estimated (Simple Asymptotic Method with Continuity Correction) with an accuracy of +/-15 %(0.65: 0.95), the number of subjects needed is 35. Taking into account 20 % of lost to follow-up and/or data that could not be used with either technique, the number of subjects to be recruited was therefore set to 42 subjects.

### Measurement methods

#### Digitized planimetry

After necessary care, including possible debridement, a transparent, non adhesive dressing (HydroTac® transparent Comfort, Hartmann) was applied on the wound. Then, the investigator draws the contours (outer edge) of the wound on a transparency superimposed on the transparent dressing. At the end of the treatment, the nursing team decides to leave the dressing in place, or to change it depending to the patient’s needs.

The area in cm^2^ is evaludated in a second time, by computer analysis of the transparent sheet with the presence of a scale, using ImageJ.

#### Woundtrack

The Woundtrack algorithm enables a semi-automatized measure of the wound surface, following a segmentation made by the clinician.

During the trial, this measure was carried out before acetate tracing, just after the necessary care. A specifically adapted sticker was put a few centimeters away from the wound, on healthy skin, in the same plane as the wound. If no site was available, it was attached on a tongue depressor and presented close to the wound, in the same plane. This sticker contains four ArUco markers. A snapshot was then taken using the Pixacare application, installed on a smartphone (iPhone 12). The image was centred on the wound, ideally at a distance of 20 to 30 cm. A light signal indicated that the ArUco markers have been recognized, and thus that the image was valid. The clinician then trimmed the wound on the smartphone.

The Woundtrack algorithm then assessed the wound surface, using the distance between the ArCuro markers to calibrate the measurement. The area was natively expressed in cm^2^.

#### Woundsize

The Woundsize algorithm aims to achieve an automatized wound surface measurement, with a segmentation of the wound by a machine learning algorithm.

The automated analysis module Woundsize was not, at the time of conducting the trial, directly integrated into the application. The analysis was therefore performed in a second step. The acquired image, previously processed according to the Woundtrack algorithm, was thus reused and processed in two stages. The first step involved automated segmentation using a deep learning algorithm of the convolutional neural network (CNN) type (see Sun [23]).

These CNNs are inspired by the architecture of the visual cortex. The initial layers, known as convolutional, are used to analyse the image using a sliding window, which scans the image point by point. They enable the detection of patterns (edges, textures, etc.). The signal is then processed by pooling layers, which extract fundamental characteristics and reduce the dimensionality of data from convolution layers. Finally, fully connected layers abstract this information and enable to distinguish the wound from healthy skin.

The algorithm used by Woundsize is a particular form of CNN, ResNet (for *deep residual network*), as described by He and al. in 2015 [10]. This architecture exploits the presence of skip connections, which avoids the degradation of results on more complex architectures and speeds up model training.

During the training phase, the algorithm was fed with images of chronic wounds, in which manual segmentation was carried out by clinicians with expertise in chronic wounds.

The second step consisted in applying a method similar to Woundtrack algorithm, allowing the measure of the segmented area.

### Population

#### Quantitative analysis of measured areas

The patients included were mainly women (24/42), with a mean age of 77,9 years. The phototype, estimated by Fizpatrick’s grading was predominantly 3 (81 %) and 2 (14.3%).

Of the 42 patients included in the study, six were not included in the analysis because their wounds did not meet the exploitability criteria (complex wounds in several planes: 4 cases; wound with difficult edge delineation, due to periwound haematoma or severe edge maceration: 2 cases). These patients were excluded without taking into account the exploratory data analysis performed.

For the 36 remaining patients, we considered wound surface measurements using three methods:

- Digitized planimetry (PL), considered as the reference method;
- Woundtrack algorithm (WT), allowing a semi automated wound area measurement, after delimitation of the lesion by the expert;
- Woundsize algorithm (WS), allowing an automated measure of the wound area. These measures were taken independently by two practitioners (Experts A and B).

Three pairs of expert-matched data sets are described: SPLA and SPLB (digitized planimetry); SWTA and SWTB (Woundtrack algorithm); SWSA and SWSB (Woundsize algorithm).

These pairs were concatenated into global groups called SPL, SWT, SWS. These data were checked for normality using Shapiro’s method.

They are summarized in tables 1, 2 and 3.

**Table 1.** Digitized planimetry - SPL (cm^2^)

|  | SPLA | SPLB | SPL |
| --- | --- | --- | --- |
| Count | 36 | 36 | 72 |
| Mean | 17.81 | 17.62 | 17.71 |
| Std | 24.72 | 24.99 | 24.69 |
| min | 2.8 | 2.21 | 2.8 |
| 25% | 5.96 | 6.11 | 5.96 |
| 50% | 10.66 | 9.17 | 9.8 |
| 75% | 19.33 | 19.91 | 19.91 |
| Max | 144.95 | 145.28 | 145.28 |
| Shapiro (p value) | < 0.001 | < 0.001 | < 0.001 |

**Table 2.** Woundtrack - SWT (cm^2^).

|  | SWTA | SWTB | SWT |
| --- | --- | --- | --- |
| Count | 36 | 36 | 72 |
| Mean | 16.05 | 15.95 | 16.00 |
| Std | 19.22 | 17.61 | 18.31 |
| Min | 2.4 | 2.4 | 2.4 |
| 25% | 5.8 | 5.7 | 5,8 |
| 50% | 11.15 | 10.35 | 10.65 |
| 75% | 19.02 | 20.65 | 19,32 |
| Max | 101 | 95.9 | 101 |
| Shapiro (p value) | < 0.001 | < 0.001 | < 0.001 |

**Table 3.**
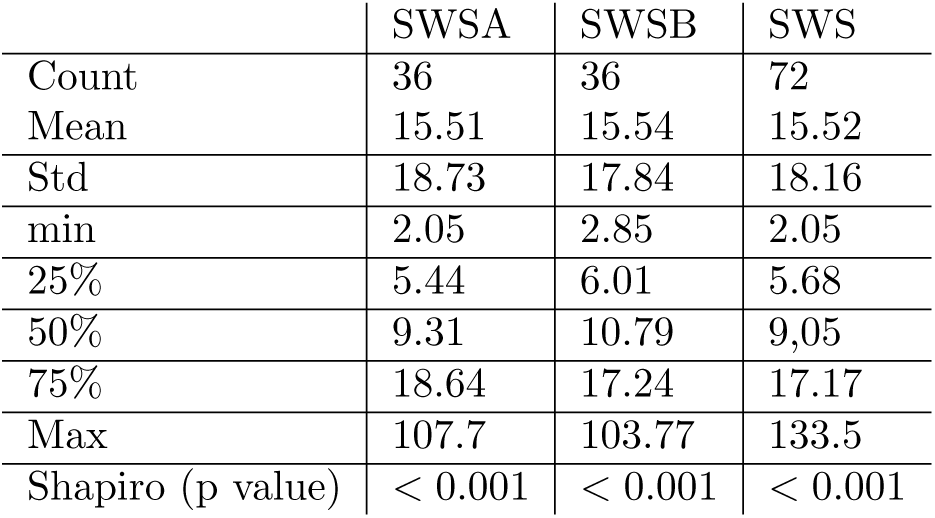
Woundsize - SWS (cm^2^).

|  | SWSA | SWSB | SWS |
| --- | --- | --- | --- |
| Count | 36 | 36 | 72 |
| Mean | 15.51 | 15.54 | 15.52 |
| Std | 18.73 | 17.84 | 18.16 |
| min | 2.05 | 2.85 | 2.05 |
| 25% | 5.44 | 6.01 | 5.68 |
| 50% | 9.31 | 10.79 | 9,05 |
| 75% | 18.64 | 17.24 | 17.17 |
| Max | 107.7 | 103.77 | 133.5 |
| Shapiro (p value) | < 0.001 | < 0.001 | < 0.001 |

### Statistical analysis

We sought to compare the methods evaluated (WoundTrack and WoundSize) with the reference method (Digitized Planimetry).

The two main aspects of the evaluation were:

- *Accuracy* : This refers to how close the measured values are to the true or accepted value. It indicates the correctness of the measurement;
- *Precision*: This refers to the consistency or reproductibility of the measurements. It indicates how close repeated measurements are to each other.

The statistical analysis will proceed through a four steps path.

*An overall analysis of the three datasets* using a multivariate analysis of variance on repeated measures (Friedmann’s test, followed by Nemenyi’s) will give us an initial overview on the agreement of the three methods.

We will refine our analysis through by evaluating of the *precision* (or consistency) of each test by studying the difference between the measurement made by the two experts, to assess the reproductibility of the test (Pearson’s correlation, percentage of shared variance, standard deviation of the differences, coefficient of variation).

Next, we will consider to evaluate the *accuracy* of the two evaluated test regarding the reference method (Student’s t-test, Pearson’s correlation, shared variance, linear regression).

A fourth step was added after the preliminary statistical analysis. Graphical analysis of the results showed the existence of a high non-conformity rate for the methods studied, defined as a deviation of more than 20% in absolute value from the reference method, on surface measurements of less than 8 *cm*^2^. We considered this error to be due to the significant impact on the surface of limited variations during segmentation. We analyzed, in this subset of measurement, *the spread of measurement* between the two experts, then the existence of a significant difference of the distribution and variance between the evaluated method and the digitized planimetry (tests of Levene, Bartlett and Wilcoxon).

The interpretation scale of the Intra-Class Correlation value was adapted from the publication of Koo and Li [13]:

- ICC values less than 0.5: poor reliability
- ICC values between 0.5 and 0.75: moderate reliability
- ICC values between 0.75 and 0.9
- ICC values greater than 0.9: excellent reliability

The statistical analysis was carried out on Python, using the libraries *pandas* for data manipulation, *matplotlib* and *seaborn* for data visualization, and *scipy* for statistical analysis.

## Results

### Multivariate analysis

As tables 1, 2 and 3 show, the distribution of the three datasets was not normal: despite an ANOVA, we perform a Friedmann’s test, completed by a post-hoc comparison by Nemenyi’s test.

Friedmann’s test showed no significant difference between the three methods (p = 0,82).

The post-hoc comparison also demonstrated the absence of significant difference between the three methods, with a p-value calculated at 0,81 ; 0,9 ; 0,87 respectively for the comparison pairs SPL-SWS; SWP-SWT; SWS-SWT.

We can conclude that the multivariate analysis of the variance didn’t objectify a significant difference between the three measurement modalities.

### Precision

We realized a paired analysis of each dataset (expert A vs B). Results of the statistical analysis are shown in the table 4.

**Table 4.** SPL, SWT, SWS: Evaluation of the precision.

|  | SWT | SWS | SPL |
| --- | --- | --- | --- |
| ICC Pearson | 0,96 ($p < 0.001$) | 0,94 ($p < 0,001$) | 1 ($p < 0.001$) |
| Percentage of shared variance | 92.53% | 88.68% | 99.69% |
| std deviation of the differences | 5.22 | 9.37 | 1.4 |
| Mean Coefficient of variation | 8.51% | 13.53% | 5,41% |

All the three methods show an excellent correlation between the measures made by two independant experts. Although SPL has near-perfect results (ICC = 1, mean coefficient of variation 5.41%), SWT shows only a slight degradation of the reliability. This is illustrated by the figure 1, representing on a Bland-Altmann graph the spread between the apparied values of SWT and SPL.

It figures:

- *An mean of differences* (solid line) that is null for both methods: no systematic bias;
- *Agreement limits* (dotted line) wider for SWT than for SPL, indicating greater variability. However, in both cases, most of the points lie between the approval limits: consistent measurements in both cases, with acceptable agreement;
- *A distribution of differences* showing greater dispersion with SWT, suggesting greater divergence of measurements with SWT.

SWS shows a greater variability of measurement, although the ICC remains excellent. A more detailed analysis of the segmentation differences obtained will be required to detail this point (see Supplementary materials: Qualitative assessment of segmentatin failure).

### Accuracy

#### Analysis of the correlation

We realized an analysis of the correlation of SWT and SWS with the value obtained via the reference method: SPL. Results of the statistical analysis are shown in the table 5.

**Figure 1.**
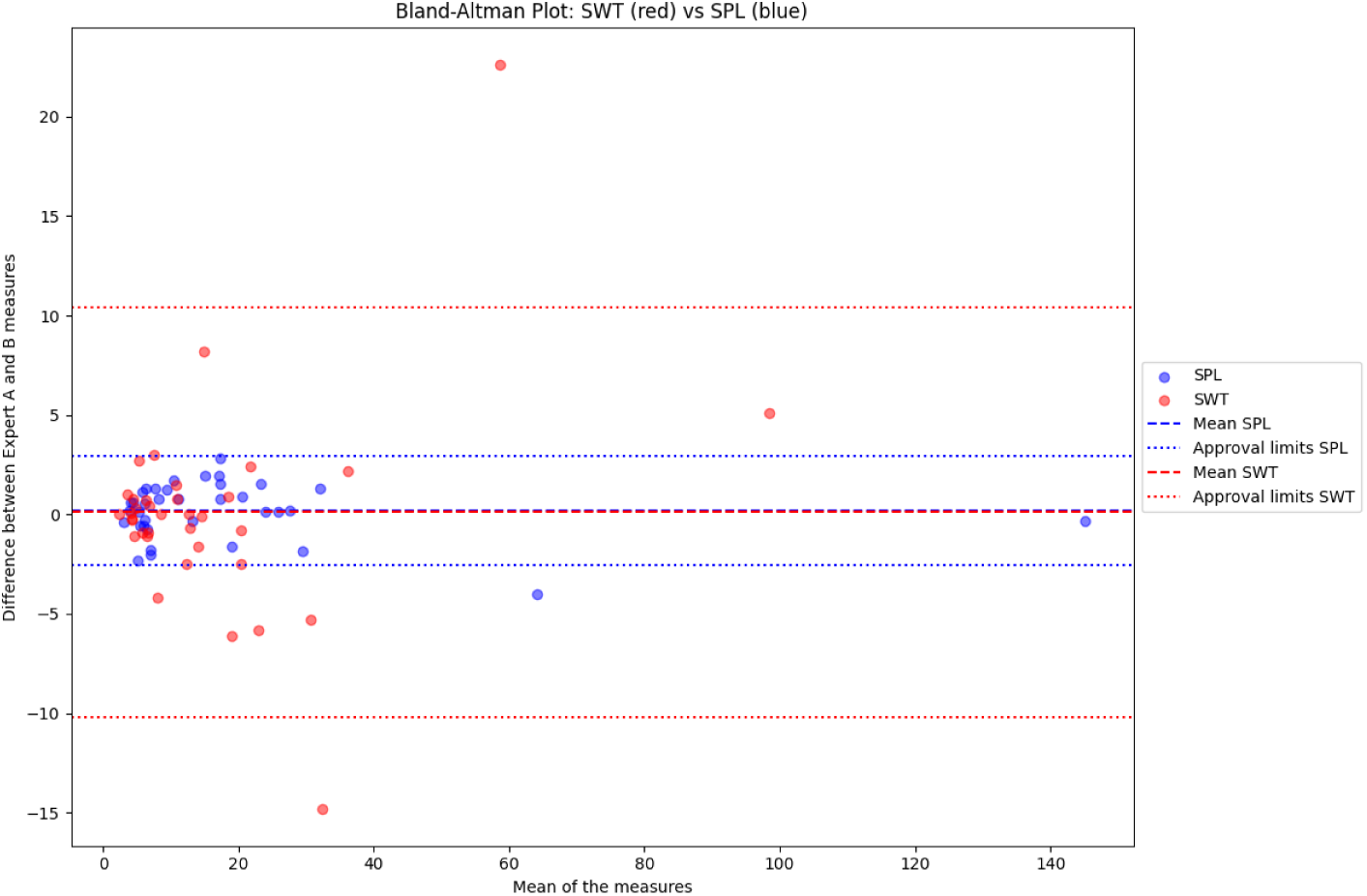
Bland-Altman plot, SPL and SWT.

**Table 5.** Correlation between SWT/SPL and SWS/SPL.

|  | SWT | SWS | SPL |
| --- | --- | --- | --- |
| Mean ($cm^2$) | 16.01 | 15.52 | 17.71 |
| t-test (p-value) | 0.64 | 0.54 |  |
| ICC (vs SPL) | 0.96 ($p < 0.001$) | 0.95 ($p < 0.001$) | |
| Percentage of shared variance | 92.56 % | 90.19 % |  |

#### SWT: linear regression of SPL by SWT

As shown in figure 2, there is a linear relationship (slope 1.3) between SWT and SPL, with close agreement between the two variables (correlation coefficient: 0.96). The standard error is 0.04. This linear regression is significant (p *<* 0.001).

#### Bland-Altmann analysis: SPL and SWT

The figure 3 shows a mean bias close to zero, with the majority of points within the limits of agreement, and a homogeneous distribution of points around the mean bias line: it suggests good agreement between SWT and SPL, with minimal mean bias and differences within the limits of agreement.

#### Linear regression of SPL by SWS

This shows a linear relationship very close to that obtained with SWT (slope 1.29; correlation coefficient 0.95, standard error 0.05, *p <* 0.001).

### Non-conformities in small wounds

Non-conformities were defined as a spread of more than 20% in absolute value of the measurement obtained by evaluated method, compared to the Digitized Planimetry.

**Figure 2.**
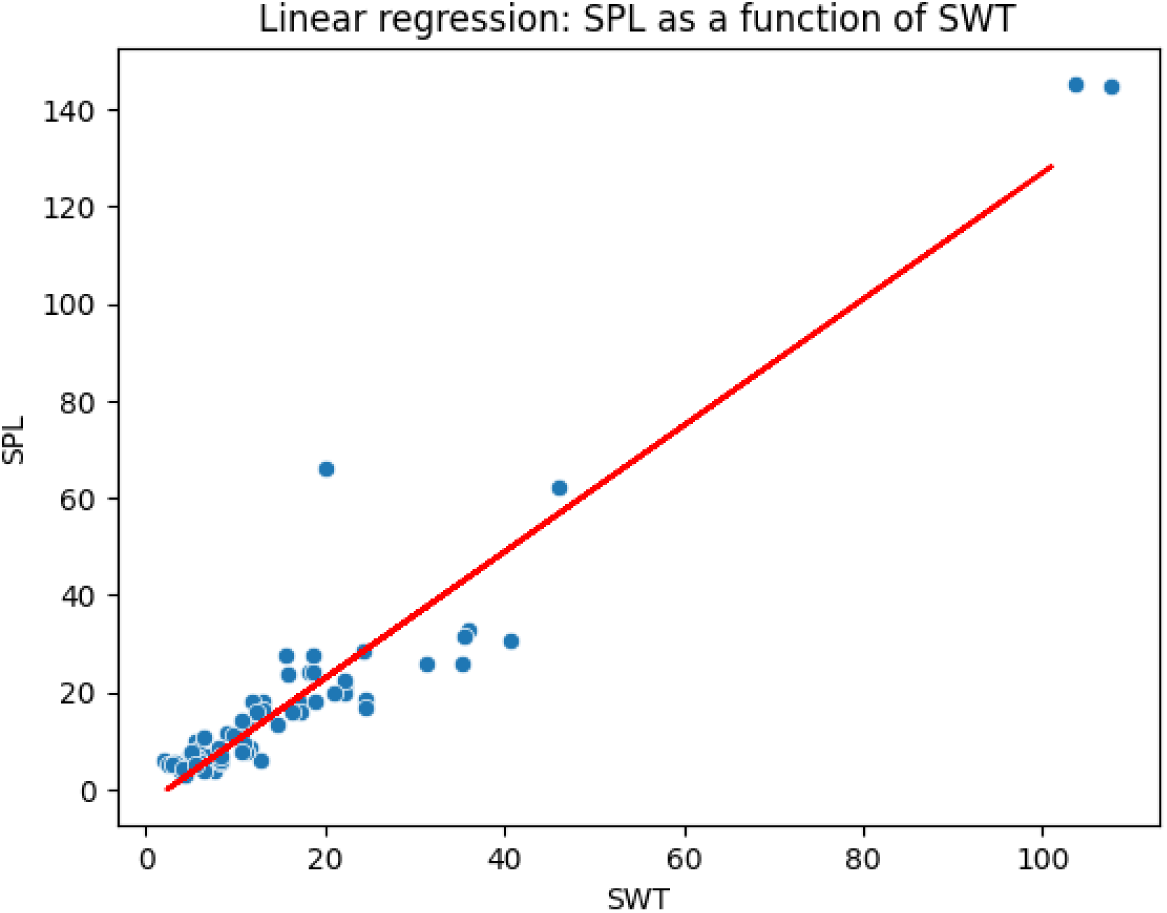
Linear regression of SPL by SWT.

#### Woundtrack non-conformities in small wounds

70.1% of the surfaces measured by Woundtrack were in conformity with the digitized planimetry.

Figure 4 shows the distribution of the differences between SWT and SPL: it reveals a peak of non-conformities in areas smaller than 8cm^2^.

We hypothesized that measurement methods based on segmentation may be too coarse to enable effective measurement of small areas, where a small difference will have a major impact when expressed in relative terms.

For example, for a disc with a diameter of 1.5cm (calculated area: 7 cm^2^), a variation of plus or minus 2 mm on the radius will give a measurement ranging from 5.3 cm^2^ to 9cm^2^, i.e. a potential variation of over 50%. However, the tracing methods used for planimetry are quite imprecise in this aspect.

We therefore set out to compare the errors made by the two methods in small areas, defined as the measurements for which SPL *≤*8 cm^2^. For each element in the subset, we considered the variable DiffSPL = SPLA- SPLB, and DiffSWT = SWTA - SWTB.

These two variables are shown in figure 5.

Both variables appear to be centered around 0, suggesting that mean differences may be close to zero in both cases. DiffSPL seems to have a more consistent distribution, with fewer extreme values.

We then compared the distribution and the variance of the two variables. Normality of the distribution were assessed for both variables by a Shapiro test (DSPL: p = 0,38 ; DSWT: p = 0.06). We then performed:

- A Levene’s test, a parametric test to compare the variance of the two variables: no difference (p = 0,62);
- A Bartlett’s test, a non-parametric test to compare the variance of the two variables: no difference (p = 0,14)
- A Wilcoxon’s test, a non-parametric test to compare the distribution of the two variables: no difference (p = 0,76).

These statistical results qualify the visual interpretation of the violin plot. Although the plot suggests visual differences, particularly in terms of dispersion and distribution shape, statistical tests indicate that these differences are not statistically significant.

**Figure 3.**
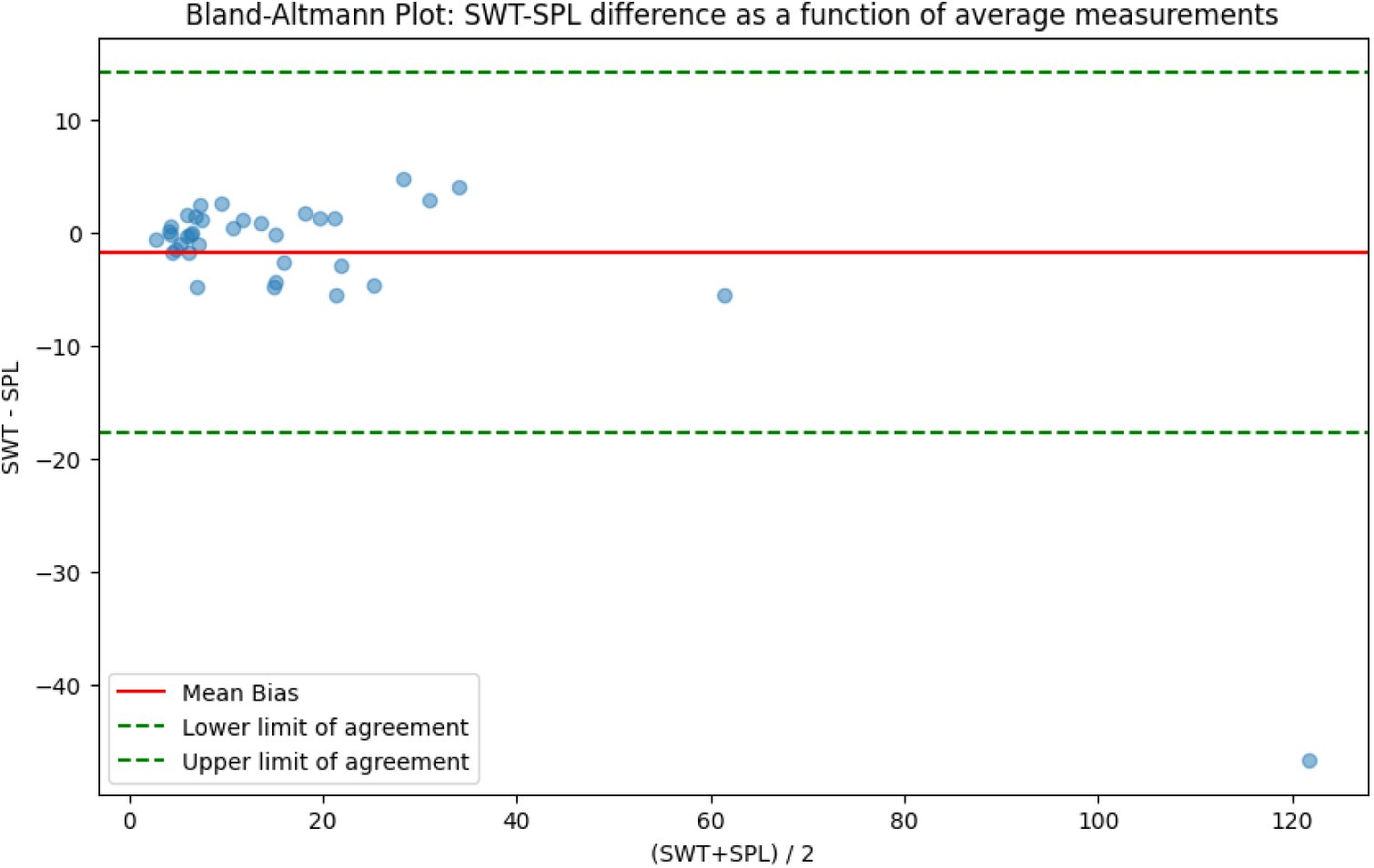
Bland-Altmann plot, difference between SWT and SPL.

#### Woundsize non-conformities in small wounds

63% of surfaces measured by Woundsize were in conformity with the digitized planimetry. We performed the same statistical analysis between SWS and SPL in the small area subset.

We represented those two variables in figure 6. The difference in dispersion between diffSWS and diffSPL is more pronounced than previously seen with diffSWT. DiffSWS shows significantly greater variability than diffSPL.

**Figure 4.**
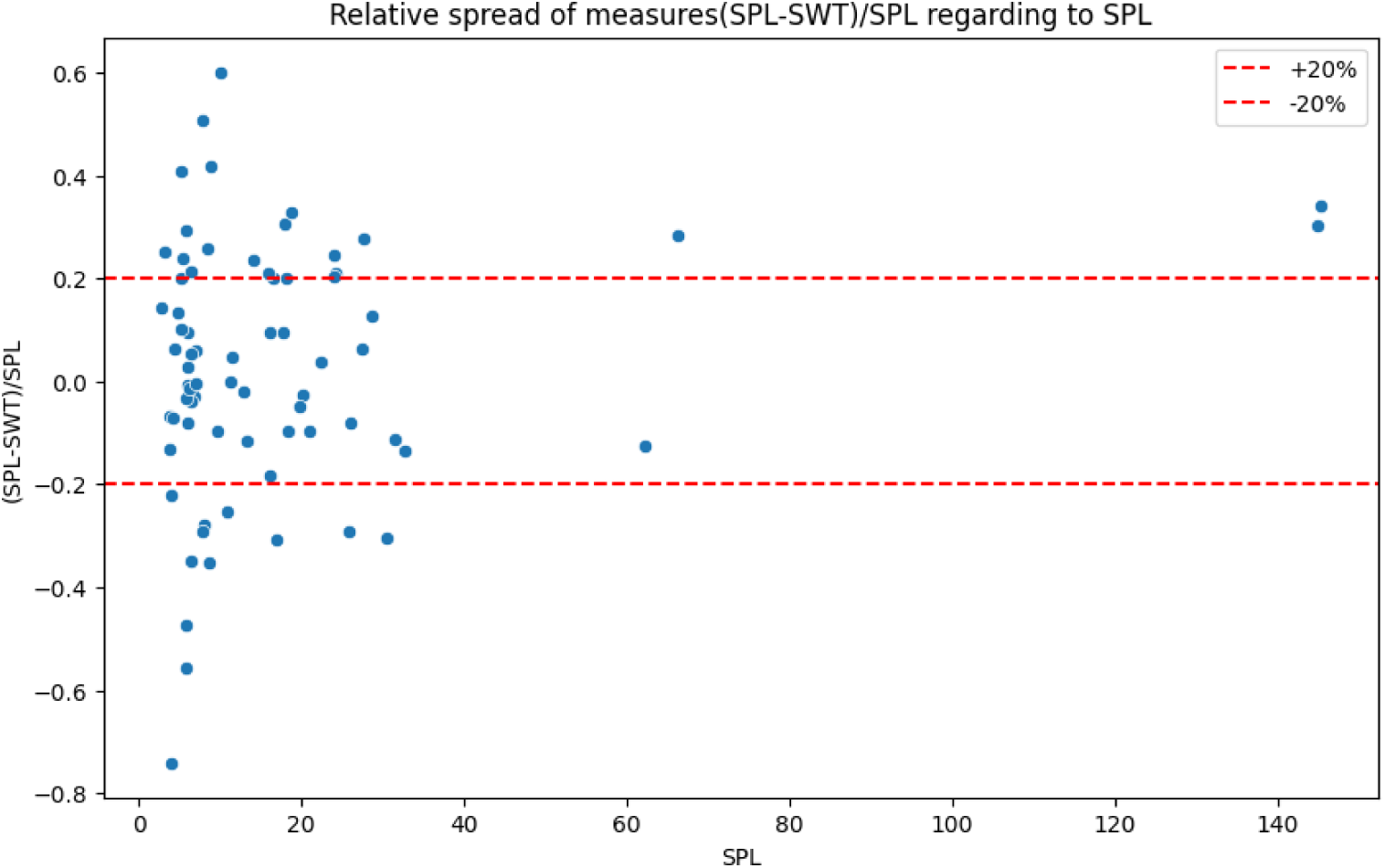
Relative variation of SWT regarding to WPL.

**Figure 5.**
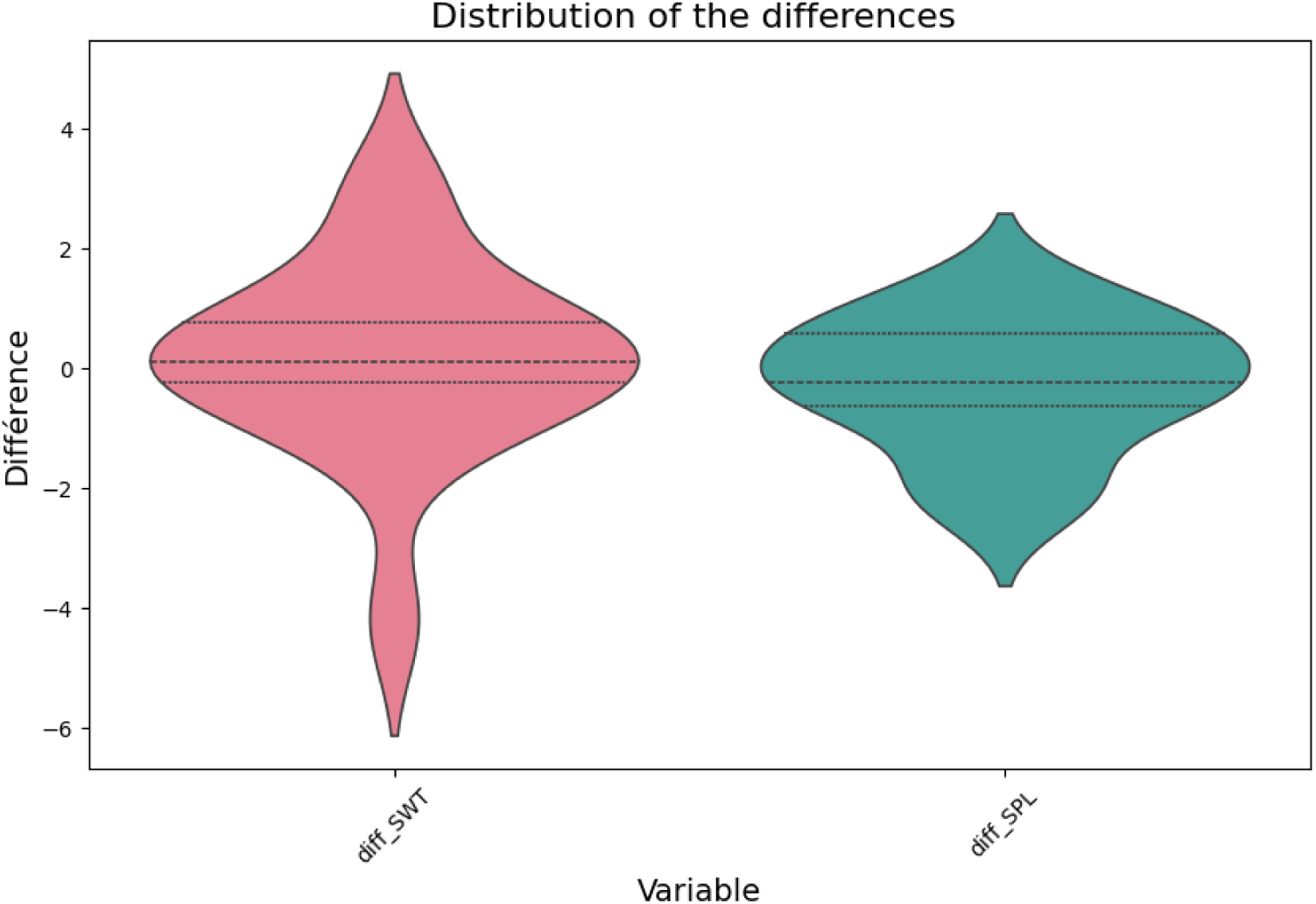
Distribution of the differences in paired measures.

**Figure 6.**
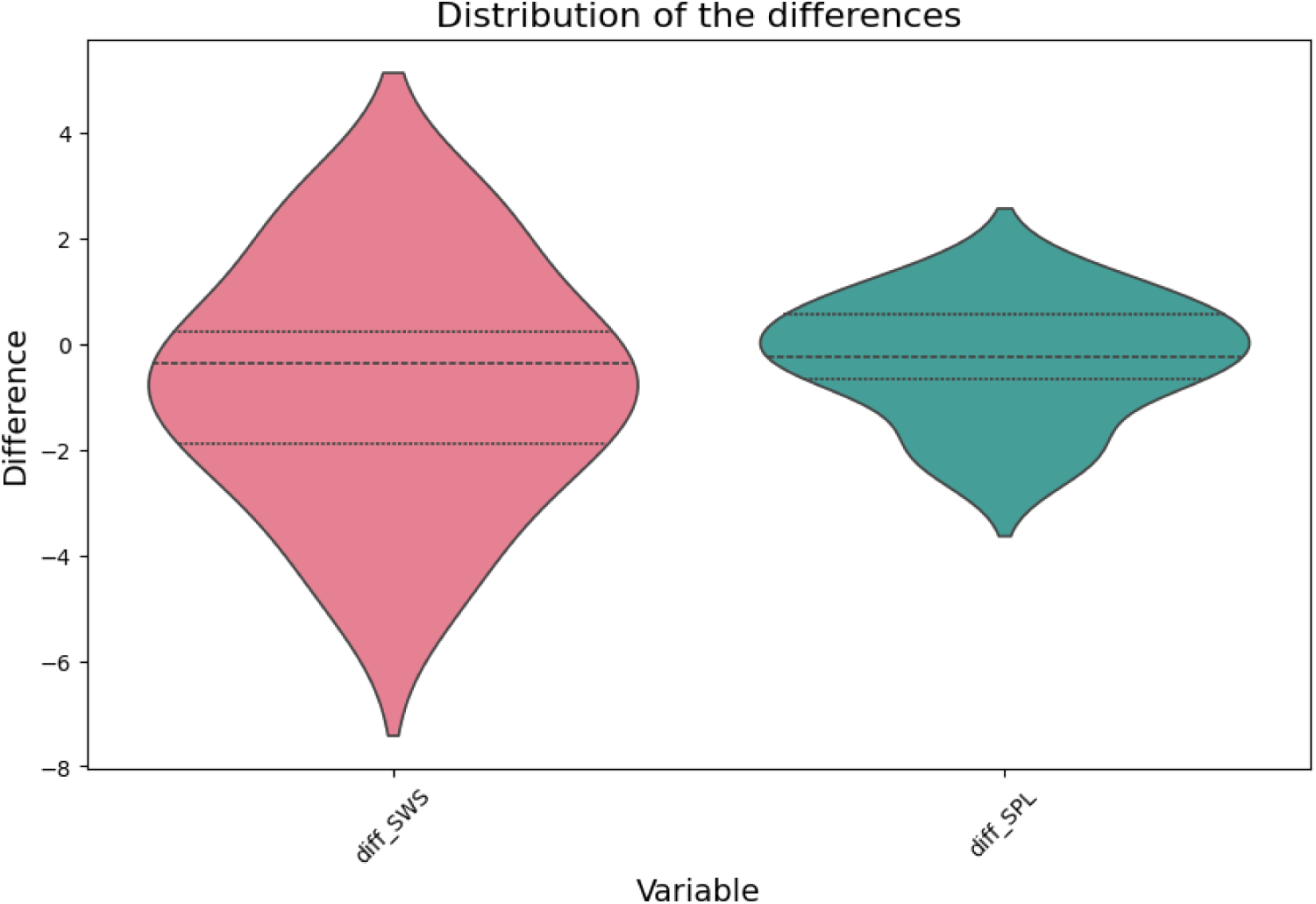
Distribution of the differences in apparied measures.

We performed the same statistical analysis as above. Shapiro’s test was used to assess the normality of the distribution (DiffSPL p = 0.38 ; DiffSWS: p = 0.70). We realized:

- Levene’s test: variances differs with statistically significance (p = 0.03).
- Bartlett’s test: variances differs with statistically significance (p = 0.01)
- Wilcoxon’s test: no difference in distribution (p = 0,27).

They confirm that DiffSWS shows significantly greater variability than DiffSPL, as suggested by the graph. However, despite this difference in dispersion, the overall distributions of the two variables do not differ significantly.

## Discussion

### Methodological consideration

In the context of evaluating the performance of algorithms for measuring the surface of chronic wounds, it is crucial to assess both the precision and accuracy of these methods. One effective methodological approach is to conduct a paired study, where each wound is measured using both the test algorithm and a reference method. This paired design enables a direct comparison of measurements obtained from the two methods, providing a robust assessment of the test algorithm’s accuracy and reliability.

#### Data collection by two experienced practitioners

made it possible to validate the data collection, whether through more reliable measurements or through greater consistency in the collection process. This approach to data collection could potentially constitute a bias, as the practitioners’ expertise could potentially modify the measurement results. However, the expertise related to conventional methods and not to the practice of smartphone measurements: while the use of Pixacare as a photo library was common practice, the measurement methods had never been used by either of the experts. We find this reflected in the data presented above. Planimetry measurements show extremely high precision, while the main pitfall of automated measurements related to the image acquisition method, particularly regarding exposure (see Supplementary material: Qualitative assessment of segmentation failure).

#### The choice of the reference method

is particularly critical. Digitized planimetry stands out as the preferred reference method due to its extensive validation and ability to measure wounds in clinical practice. This method provides direct, real-time measurements without the distortions inherent in photographic methods. In contrast, studies relying on photographic measurements introduce potential biases and inaccuracies due to variations in lighting, angle, and lens distortions. These issues can significantly compromise the reliability and validity of the results.

Therefore, the selection of digitized planimetry as the reference method ensures the robustness and clinical relevance of the performance evaluation. This approach not only enhances the credibility of the findings but also provides a more accurate basis for comparing the test algorithm’s effectiveness in real-world clinical settings.

### Results of the study

The multivariate analysis performed showed that the measurement obtained with the three methods didn’t differ in term of variances. Subsequent analysis enabled us to consider the method with more details.

#### Woundtrack is comparable to the digitized planimetry

We demonstrated that the *precision* of Woundtrack is very close to digitized planimetry, which showed exceptional results in our trial: we can consider that Woundtrack is effective to realize longitudinal follow-up of wound area.

Assessment of the *accuracy* of the Woundtrack algorithm is similar, with a close correlation between WT and PL measurements. We can consider that Woundtrack provides a measurement of the area of the chronic wound as reliable as digitized planimetry.

The analysis of the non-conformities in the small-area subset showed that the differences between paired measures realized by PL ou WT were not significantly different - measurements taken by different operators are as reproducible in one method as in the other. The underlying mechanism, corresponding in our hypothesis to an offset in the manual segmentation process, affects both methods in the same way. We have to consider that the absolute difference in the measured area in this subset is very small, and has no impact on the values.

Improving the process is likely to be difficult because we also need to consider a major difference between the two techniques, with regard to wound geometry.

Full-contact methods, including digitized planimetry, apply a deformable sheet to the wound, thereby respecting its geometry and dimensions. Partial-contact’s methods, such as Woundtrack, work on the basis of a photo, which involves projecting a complex geometry onto a plane, and thus working on a deformed image. In order to evaluate this effect, we have (see below, Supplementary materials: Woundtrack on transparent tracing.) measured the surfaces of the tracings used for planimetry with WT algorithm. We observed an improvement in the results, particularly for the measurements of the largest wounds, with correlation indices of 0.99.

This limitation is inherent in all partial contact methods, and must be taken into account for the largest wounds. The method described ‘Wountrack on transparent tracing’ provides an answer to this limitation, and is also applicable to the most complex wounds, located in several planes.

Considering the ease and the speed with which this algorithm can be implemented compared to digitized planimetry, it seems to us that the Woundtrack algorithm can be considered as equivalent to the reference method for measuring the surfrace of chronic wound, not least because the possibility of multiplying measurements makes it possible to compensate for marginal differences.

#### WoundSize results are lower than the others two

The satisfactory results obtained in the multivariate analysis for the Woundsize algorithm were not confirmed by an analysis comparing it directly with digitized planimetry.

**Table 6.** Comparison of relaibilities between partial contact mobile application.

| Author | Device | Inter-rater reliability | Intra-rater reliability |
| --- | --- | --- | --- |
|  | Woundtrack | 0.96 | 0.96 |
|  | Woundsize | 0.94 | 0.95 |
| Fong [8] | TA | 0.701 - 0.864 | 0.977-0.984 |
| Biagioni [2] | Imito | 0.978 | - |
| Do Khac [7] | Imito | 0.98 | 0.99 |
| Kuang [14] | NDKare | 0.98 | 0.991 |

The precision and accuracy of the algorithm showed a deviation from digitized planimetry. Analysis of non-conformities on small-area measurements also demonstrated a significant difference in the dispersion of measured values.

To explore these limitations, we carried out a qualitative analysis of segmentation failures (see Supplementary materials: Qualitative assessment of segmentation failure). 76% of the images reprocessed by Woundsize did not show any major segmentation failures. The errors found were generally major. They enabled us to understand the algorithm’s methods (edge zone search, corresponding to a zone of high contrast, with a threshold in the thickness of the associated healthy skin zone). Analysis of the causes of error showed that foreign bodies (osteosynthesis plates, haematomas) in the wound and wound complexity (hyperkeratosis, perilesional maceration) were not statistically linked to the occurrence of errors. The fundamental issue was exposure defects (overexposure from flash use, underexposure) which reduced the contrast between perilesional skin and wound. Image exposure quality was therefore the determining factor in segmentation failure. These elements lead us to consider that the method’s limitation isn’t with the algorithm. Improving it won’t necessarily lead to significant progress. We propose not using it autonomously, but integrating it into a collaborative process: Woundsize: Proposes segmentation; then Clinician: corrects and validates segmentation.

### Comparison of WT and WS with other available contact methods

The results reported by Sugama cite[sugama2007] show excellent results for the Visitrak method, both in terms of accuracy and precision. These results are obvious, as this method is very similar to acetate tracing, with the measuring equipment in contact with the wound. However, this is the real limitation of the technique, with the risk of further damage of the wound, but also the difficulty of exporting the method beyond expert centers.

### Comparison of WT and WS with other partial contact methods

Wu [26] compared the different methods (Tissue Analytics, Imito, NDKare: mobile applications and VeVMD, ImageJ, AutoCAD) accross their intra-rater and inter-rater reliability, respectively corresponding to the precision and the accuracy of the measurement. We summarized these data in the table 6, including the results ofWoundtrack and Woundsize.

These elements show that partial-contact methods give nearly-comparable results to acetate tracing or digitized planimetry in all cases: all method (at the exception of Tissue Analysics) obtained excellent (ICC *>* 0.90) intra-rater and inter-rater correlation - even Woundsize, to be less-efficient that Woundtrack.

**Table 7.** Comparison of reliabilities between no-contact measure systems.

| Author | Device | Inter-rater reliability | Intra-rater reliability |
| --- | --- | --- | --- |
| Chan [6] | C4W | 0.872-0.932 | 0.984-0.994 |
| Chan [5] | WA | 0.932-0.950 | 0.99 - 0,992 |
| Lasschuit [16] | 3DWM | 0.97 | 0.96 |
| Pena [19] | 3DWM | 0.978 | 0.988 |
| Toygar [25] | 3DWM | 0.996 | 0.997 |
| Jorgensen [12] | 3DWM | 0.996 | 0.997 |

#### Comparaison of WT and WS with other available non-contact methods

3D no-contact methods for measuring chronic wounds offer several significant advantages. Firstly, they enable accurate wound assessment without the need for markers, reducing the risk of infection and improving patient comfort. Secondly, these techniques enable a detailed evaluation of the wound volume, offering a more complete understanding of its progression. Finally, they are particularly suited to the management of complex wounds due to their ability to capture fine anatomical details even in complex geometry.

Wu [26] compared different methods, such as CARES4-WOUND (C4W), WA imaging system (WA), 3D WAM Camera (3DWM). Data comparing those devices are presented in table 7.

They show excellent results, despite the absence of skin markers. However, the use of these methods requires specific and expensive equipment, making them often inaccessible for home-based monitoring.

A hybrid approach, as proposed by Sanchez-Jimenez [24], combines 3D modeling using structure-from-motion algorithm and reconstruction of the wound with Neural Enhanced Radiance Field methods, and then the creation of an *orthoview*, which corresponds to a picture taken orthogonally to the plane of the wound . This method can be used to compare the wound progression by projection with this orthoview, offering a practical and effective solution for monitoring chronic wounds, without the need of specific skills or equipment.

### Limits of the study

#### Size of the wound

The study design considered inclusion of wounds measuring from 4 to 140 cm^2^. We found greater relative variability for wounds between 4 and 8 cm^2^. However, variations in absolute values were modest, and this dispersion of measurements was comparable to that found for digitized planimetry.

The study of larger wounds, over 60 cm^2^, is a delicate task, given the rarity of the corresponding lesions. These measurements must be taken with caution, due to the accumulation of geometric deformation. These wounds are mostly assimilated to non-developable curved surfaces (hyperbolic when “hollow”, or elliptical when ”humped”). What’s more, the quality of the lenses used in smartphone photographic sensors can create ’field curvature aberrations’ (images projected onto a curved surface) or ’geometric distortions’ (pinching or, on the contrary, swelling of the image) at the edge of the field, on an image that will be ’out of focus’. The problem is made even more complex by the image post-processing algorithms built into smartphones.

#### Phototypes represented in the study

The major limitation of our study concerns the recruitment of patients with high phototypes (5 and 6), who are absent from our study. The impact on semi-automated surface measurement (Woundtrack) is theoretically limited, as trimming is performed by the operator, and pigmentation by definition only affects epidermal areas. However, it is essential to verify this assertion. On the other hand, the impact on the automated method (Woundsize) is a risk that has to be specifically evaluated.

## Conclusion

### Woundtrack

(semi-automatized wound measurement, with segmentation by the clinician) is comparable to digitized planimetry, considered as the gold standard of the methods of measurement of the wound surface. The differences observed in the smallest wound are not significant. However, we have to be cautious in the taller wound, regarding to the complex, curved geometry of this lesions. Finally, WT results are comparable to other partial contact methods: they may be considered as good as digitized planimetry or acetate tracing in clinical pratice.

Ease of use should enable surface measurement to be used more frequently as part of wound diagnosis and monitoring. There are many benefits to be gained from this ease of quantifying wound kinetics, whether in terms of more appropriate use of dressings and invasive therapy.

### Woundsize

(fully automatized wound measurement) showed good (accuracy) or excellent (precision) results, but its results are poorer than those of WT, especially in small wounds. The cause of the failure was mainly represented by the quality of the input images (over- or under- exposure). Technical optimization of the process may be deceptive. Integrating WS in a collaborative workflow (algorithm: propose then Clinican: correct and validate) seems to be the most efficient way to optimize the process.

## Data Availability

Available on request to the corresponding author

## Supplementary materials: Woundtrack on transparent tracing

As seen *supra*, Woundtrack offers an effective, simple, partial contact method for mesauring the wound surface. However, there is a slight difference with the reference method, which we consider to be due to the segmentation process. To clarify the impact of the segmentation method (tracing on transparency versus tracing on the smartphone screen), and of the geometrical complexity of the wound (the image is a projection of a curved surface onto a flat detector), we measured the area of the acetate tracing with the Woundtrack algorithm, to compare it to the measure obtained in planimetry.

### Population and method

We considered the same population as *supra*. We carried out a complementary measurement of transparent tracings made with the Woundtrack algorithm. We obtained two paired measures of the 36 wounds studied. The data were named SPLWTA, SPLWTB, and SWPWT. Those datas are summarized in table 8.

**Table 8.** Woundtrack on transparent (cm^2^).

|  | SWPLWTA | SPLWTB | SPLWT |
| --- | --- | --- | --- |
| Count | 36 | 36 | 72 |
| Mean | 18.1 | 16.61 | 17.35 |
| Std | 25.39 | 23.43 | 24.27 |
| min | 3.2 | 2.3 | 2.3 |
| 25% | 5.67 | 5,6 | 5.67 |
| 50% | 9 | 9.05 | 9 |
| 75% | 20.12 | 17.17 | 19.5 |
| Max | 142.4 | 133.5 | 142.4 |
| Shapiro (p value) | < 0.001 | < 0.001 | < 0.001 |

We carried out a statistical analysis with the same framework as above, considering evaluation of the test *precision* (concordance between the two experts) and the *accuracy* of the measurement (concordance with the digitized planimetry).

**Table 9.** SPL, SPLWT: Evaluation of the reliability.

|  | SPLWT | SPL |
| --- | --- | --- |
| ICC Pearson | 0.99 (p < 0,001) | 1 (p < 0.001) |
| Percentage of shared variance | 98.37% | 99.69% |
| std deviation of the differences | 4.79 | 1.4 |

### Results

#### Precision of the test

We performed a paired analysis of each dataset (expert A vs B). The results of the statistical analysis are presented in table 9.

#### Accuracy of the test

The correlation analysis between SPLWT and SPL consisted in:

- Student’s t-test: no significative difference (p = 0.63);
- Pearson’s ICC: 0,98;
- Percentage of shared variance: 92.86%.

The linear regression realized was statistically significant (p *<* 0.001) with a slope of 0.99, a standard error of 0,023.

#### Non-conformities

Non-conformities, as defined above, was only found in 7 cases, corresponding to 90.3% of conformity. Here too, most non-conformities were found on small lesions.

Non-conformities observed in larger lesions are no longer found.

### Conclusion

This additional analysis supports the study’s conclusions. The optimization of the correlation, especially in large wounds, emphasizes the importance of the curvature of the wound, that led to a deformation during its projection on a flat detector, whereas the use of transparent tracing applies a deformable layer that gives a better conformation to the curvature.

It also makes possible to define a simplified method of digital planimetry, that eases the acquisition procedure and enabling it to be carried out rapidly, extemporaneously. The procedure would consist of:

- Clean and even debride the wound
- Place a transparent dressing over the wound, mark out the wound
- Place the dressing on a white sheet, stick on a sticker, measure the wound with Woundtrack

This would reduce the acquisition procedure by one step (interposing an element between the transparency and the wound for hygiene reasons) and enable immediate measurement, without going through the digitization and ImageJ analysis stages.

This would make it possible to overcome the current limitations of measurement on photographic plates, whether for large wounds or multiplanar wounds.

## Supplementary material: Qualitative assessment of segmentation failures

Analysis of the data collected showed that there were significant differences between the values collected by the two algorithms, Woundtrack and Woundsize. As the surface measurement method is identical, the segmentation process is the element in question. We therefore carried out an evaluation of the errors and looked for their causes, by means of a qualitative analysis.

### Materials

The raw, unsegmented images were re-used. Only matched images were used, and two images for which one of the two elements was missing were excluded from the study. One pair of images for which segmentation was deemed impossible (presence of significant maceration of the edges, preventing satisfactory delineation) was excluded. Work was therefore carried out on 38 pairs of images, taken independently by experts A and B.

### Methods

The analysis was carried out in two stages.

A first operator (GM) analyzed the segmented images and discriminated between them according to the quality of the segmentation: a segment was considered correct if the sum of the false-positive areas (area of healthy skin included in the segmentation) and false-negative areas (area of the wound excluded from the segmentation) was less than 10%.

This analysis of the images enabled us to understand how the algorithm worked, using the contrast between healthy skin and the wound over a threshold thickness to determine the edge of the wound. Based on this observation, an analysis of the pairs of images for which the segmentation was inconsistent identified several criteria that could explain the segmentation errors. Difficulty in manually trimming the wound was the first overall criterion. Two specific criteria were also considered, firstly the complexity of the wound edge (hyperkeratosis, maceration and ulceration of the periwound skin), and secondly defects in the exposure of the images (first and foremost, underexposure darkening a portion of the image, but also, more marginally, overexposure when the flash was used). A fourth criterion, which seemed less relevant, was added: the existence of foreign bodies (haematomas, osteosynthesis plates).

In the second stage, a second operator (TM) blindly analyzed the images, in pairs, by characterizing them according to the 4 criteria mentioned above: difficult segmentation, irregular margins, exposure defect and presence of foreign bodies.

A univariate statistical analysis was performed, after drawing up contingency tables, by calculating *χ*^2^. A second multivariate analysis was then performed, using logistic regression. These analyses were performed in Python, using the scipy.stat and sklearn.linear-model libraries.

### Résultats

18 of the 76 images studied (23.6%) showed a segmentation defect. Both paired images were affected in 5 cases (5/38 pairs, 13%).

Univariate analysis showed a significant association between exposure defects (*χ*^2^=17.5, p*<* 0.001), while the other explanatory variables were not statistically significant, either the complex wound (*χ*^2^= 2.7095, p = 0.0998) or the other criteria,

Multivariate analysis using logistic regression confirmed these relationships after adjustment for all variables. Lack of exposure appeared to be the strongest predictor of segmentation defects (OR=19.15, CI95% [3.91-93.82], p*<* 0.001). Wound complexity showed a non-significant trend (OR=3.42, CI95% [0.55-21.20], p=0.19). The presence of a segmentation difficulty identified by the expert was not associated with effective segmentation defects (OR=1.24, CI95% [0.24-6.35], p=0.79). Foreign bodies had no significant influence on segmentation quality (p=0.99).

## Discussion

The convergence of univariate and multivariate analyses shows the importance of the quality of exposure of the wound when the images are taken. Both over- and under-exposure led to a reduction in contrast between healthy skin and wound, which prevents effective delineation. The complexity of the wound (hyperkeratosis, macerated wound with dermabrasion around the edge, for example) and the presence of a foreign body (osteosynthesis plate, haematoma in the wound) do not play a significant role in these failures.

If the determinant of segmentation quality is the quality of the image, this means that the limiting factor in automated segmentation has more to do with the quality of the collection than with the robustness of the algorithm. The improvement in its discrimination capacity will not be sufficient to compensate for variations in the quality of input images. This is all the more important given that the study was carried out using standardised equipment (iPhone 12), and that data collection in everyday practice will be highly variable both immediately (variations in the skills of the carers taking the images, variations in the quality of the sensors used) and over time (improvements in sensors and software post-processing by smartphones, acquisition of experience).

It is therefore probably impossible under these conditions to propose an autonomous system, operating independently of human control. It would be much more appropriate to use it as a decision-making tool, integrated into a collaborative process such as Algorithm: Proposal - Clinician: Correction then Validation.

## Supplementary materials: acquisition of the sequences in Pixacare

link toward the explanatory video. QR code

## Author’s contribution

All authors contributed to the reviewing of the study.

Data collection (Pixaire-1): C.Mori, G.Maxant

Data collection (Supplementary materials): T.Maxant

Data curation, exploratory data analysis: G.Maxant, M.Pastrav

Statistical analyis: C.Bertaux

Redaction: G.Maxant

The authors would like to thank Vincent Marceddu, CTO at Pixacare, to his contribution concerning the technical aspect of Woundtrack and Woundsize processes.

## Notes

### Competing Interest Statement

The authors have declared no competing interest.

### Clinical Trial

NCT05846152

### Funding Statement

No funding received

### Author Declarations

ID clinical trial.gov: NCT 05846152 ID French CPP registration: N ID RCB 2022-A011546-37 date of ANSM autorisation: 2/9/22. Date of CPP comity favorable autorisation: CPP Nord Ouest III, 21/10/2022

## References

1. L. Barnsbee, Q. Cheng, R. Tulleners, X. Lee, D. Brain, and R. Pacella. Measuring costs and quality of life for venous leg ulcers. International Wound Journal, 16(1):112–121, 2019. eprint: https://onlinelibrary.wiley.com/doi/pdf/10.1111/iwj.13000.

2. R. B. Biagioni, B. V. Carvalho, R. Manzioni, M. F. Matielo, F. C. Brochado Neto, and R. Sacilotto. Smartphone application for wound area measurement in clinical practice. J Vasc Surg Cases Innov Tech, 7(2):258–261, June 2021.

3. M. Bilgin and U. Y. Günes. A comparison of 3 wound measurement techniques: effects of pressure ulcer size and shape. *Journal of wound, ostomy, and continence nursing : official publication of The Wound*, Ostomy and Continence Nurses Society, 40(6):590–593, 2013.

4. M. Cardinal, D. E. Eisenbud, T. Phillips, and K. Harding. Early healing rates and wound area measurements are reliable predictors of later complete wound closure. Wound repair and regeneration : official publication of the Wound Healing Society [and] the European Tissue Repair Society, 16(1):19–22, 2008.

5. K. S. Chan, Y. M. Chan, A. H. M. Tan, S. Liang, Y. T. Cho, Q. Hong, E. Yong, L. R. C. Chong, L. Zhang, G. W. L. Tan, S. Chandrasekar, and Z. J. Lo. Clinical validation of an artificial intelligence-enabled wound imaging mobile application in diabetic foot ulcers. International Wound Journal, 19(1):114–124, 2022. eprint: https://onlinelibrary.wiley.com/doi/pdf/10.1111/iwj.13603.

6. K. S. Chan and Z. J. Lo. Wound assessment, imaging and monitoring systems in diabetic foot ulcers: A systematic review. International Wound Journal, 17(6):1909–1923, 2020. eprint: https://onlinelibrary.wiley.com/doi/pdf/10.1111/iwj.13481.

7. A. Do Khac, C. Jourdan, S. Fazilleau, C. Palayer, I. Laffont, A. Dupeyron, S. Verdun, and A. Gelis. mHealth App for Pressure Ulcer Wound Assessment in Patients With Spinal Cord Injury: Clinical Validation Study. JMIR Mhealth Uhealth, 9(2):e26443, Feb. 2021.

8. K. Y. Fong, T. P. Lai, K. S. Chan, I. J. L. See, C. C. Goh, S. Muthuveerappa, A. H. Tan, S. Liang, and Z. J. Lo. Clinical validation of a smartphone application for automated wound measurement in patients with venous leg ulcers. International Wound Journal, 20(3):751–760, 2023. eprint: https://onlinelibrary.wiley.com/doi/pdf/10.1111/iwj.13918.

9. G. Gethin and S. Cowman. Wound measurement comparing the use of acetate tracings and VisitrakTM digital planimetry. Journal of Clinical Nursing, 15(4):422–427, 2006. eprint: https://onlinelibrary.wiley.com/doi/pdf/10.1111/j.1365-2702.2006.01364.x.

10. K. He, X. Zhang, S. Ren, and J. Sun. Deep Residual Learning for Image Recognition, Dec. 2015. arXiv:1512.03385 [cs].

11. W. J. Jeffcoate, A. J. Musgrove, and N. B. Lincoln. Using image J to document healing in ulcers of the foot in diabetes. Int Wound J, 14(6):1137–1139, Dec. 2017.

12. L. B. Jørgensen, J. A. Sørensen, G. B. Jemec, and K. B. Yderstræde. Methods to assess area and volume of wounds – a systematic review. International Wound Journal, 13(4):540–553, 2016. eprint: https://onlinelibrary.wiley.com/doi/pdf/10.1111/iwj.12472.

13. T. K. Koo and M. Y. Li. A Guideline of Selecting and Reporting Intraclass Correlation Coefficients for Reliability Research. J Chiropr Med, 15(2):155–163, June 2016.

14. B. Kuang, G. Pena, Z. Szpak, S. Edwards, R. Battersby, P. Cowled, J. Dawson, and R. Fitridge. Assessment of a smartphone-based application for diabetic foot ulcer measurement. Wound Repair Regeneration, 29(3):460–465, May 2021.

15. K. M. Lagan, A. E. Dusoir, S. M. McDonough, and G. D. Baxter. Wound measurement: the comparative reliability of direct versus photographic tracings analyzed by planimetry versus digitizing techniques. Archives of physical medicine and rehabilitation, 81(8):1110–1116, Aug. 2000.

16. J. W. J. Lasschuit, J. Featherston, and K. T. T. Tonks. Reliability of a Three-Dimensional Wound Camera and Correlation With Routine Ruler Measurement in Diabetes-Related Foot Ulceration. J Diabetes Sci Technol, 15(6):1361–1367, Nov. 2021.

17. Z. Li, F. Lin, L. Thalib, and W. Chaboyer. Global prevalence and incidence of pressure injuries in hospitalised adult patients: A systematic review and meta-analysis. Int J Nurs Stud, 105:103546, May 2020.

18. G. Maxant, M. Pastrav, I. Gogeneata, C. Bacjz, and A.-C. Bertaux. Clinical and medico-economic benefits of remote monitoring of chronic wounds. International Wound Journal, 22(1), 2025.

19. G. Pena, B. Kuang, Z. Szpak, P. Cowled, J. Dawson, and R. Fitridge. Evaluation of a Novel Three-Dimensional Wound Measurement Device for Assessment of Diabetic Foot Ulcers. Adv Wound Care (New Rochelle), 9(11):623–631, Nov. 2020.

20. M. Romanelli, V. Dini, L. C. Rogers, C. E. Hammond, and M. A. Nixon. Clinical evaluation of a wound measurement and documentation system. Wounds, 20(9):258–264, Sept. 2008.

21. C. Schneider, S. Stratman, and R. S. Kirsner. Lower Extremity Ulcers. Med Clin North Am, 105(4):663–679, July 2021.

22. J. Sugama, Y. Matsui, H. Sanada, C. Konya, M. Okuwa, and A. Kitagawa. A study of the efficiency and convenience of an advanced portable Wound Measurement System (VISITRAK). Journal of clinical nursing, 16(7):1265–1269, July 2007.

23. Y. Sun, W. Lou, W. Ma, F. Zhao, and Z. Su. Convolution Neural Network with Coordinate Attention for Real-Time Wound Segmentation and Automatic Wound Assessment. Healthcare (Basel), 11(9):1205, Apr. 2023.

24. D. Sánchez-Jiménez, F. F. Buchón-Moragues, B. Escutia-Muñoz, and R. Botella-Estrada. SfM-3DULC: Reliability of a new 3D wound measurement procedure and its accuracy in projected area. Int Wound J, 19(1):44–51, Jan. 2022.

25. I. Toygar, I. Y. Simsir, and S. Cetinkalp. Evaluation of three different techniques for measuring wound area in diabetic foot ulcers: a reproducibility study. J Wound Care, 29(9):518–524, Sept. 2020.

26. Y. Wu, L. Wu, and M. Yu. The clinical value of intelligent wound measurement devices in patients with chronic wounds: A scoping review. International Wound Journal, 21(3):e14843, 2024. eprint: https://onlinelibrary.wiley.com/doi/pdf/10.1111/iwj.14843.

27. P. Zhang, J. Lu, Y. Jing, S. Tang, D. Zhu, and Y. Bi. Global epidemiology of diabetic foot ulceration: a systematic review and meta-analysis †. Ann Med, 49(2):106–116, Mar. 2017.

